# Differential circulating proteomic responses associated with ancestry during severe COVID-19 infection

**DOI:** 10.1101/2024.07.09.24310087

**Authors:** Thomas M Zheng, Yann Ilboudo, Tianyuan Lu, Guillaume Butler-Laporte, Tomoko Nakanishi, David Morrison, Darin Adra, Lena Cuddeback, J. Brent Richards

## Abstract

**Background:** COVID-19 led to a disruption in nearly all aspects of society, yet these impacts were not the same across populations. During the pandemic, it became apparent that ancestry was associated with COVID-19 severity and morbidity, such that individuals of African descent tended to have worse outcomes than other populations. One factor that may influence COVID-19 outcomes is the circulating proteomic response to infection. This study examines how different ancestries had differential circulating protein levels in response to severe COVID-19 infection.

**Methods:** 4,979 circulating proteins from 1,272 samples were measured using the SomaScan platform. We used a linear mixed model to assess the ancestry-specific association between the level of each protein and severe COVID-19 illness, accounting for sex, age, and days since symptom onset. We then compared each ancestry-specific effect size of severe COVID-19 illness on protein level to one another in a pairwise manner to generate Z-scores. These Z-scores were then converted into p-values and corrected for multiple comparisons using a Benjamini-Hochberg false discovery rate of 5%.

**Results:** Comparing ancestries, we found that 62% of the tested proteins are associated with severe COVID-19 in European-ancestry individuals, compared to controls. We found that 45% and 22% of the tested proteins were different between COVID-19 infected and control individuals in people of African and East Asian ancestry, respectively. There was a strong correlation in effect size between ancestries. We found that individuals of European and African ancestry had the most similar response with a Pearson correlation of 0.868, 95% CI [0.861, 0.875] while European and East Asian ancestries had a Pearson correlation of 0.645, 95% CI [0.628, 0.661] and, East Asian and African ancestries had a Pearson correlation of 0.709, 95% CI [0.695, 0.722]. However, we found 39 unique proteins that responded differently (FDR < 0.05) between the three ancestries.

**Conclusions:** Examining 4,979 protein levels in 1,272 samples, we identified that the majority of measured proteins had similar responses to infection across individuals of European, African and East Asian ancestry. However, there were 39 proteins that may have a differential response to infection, when stratified by ancestry. These proteins could be investigated to assess whether they explain the differences in observed severity of COVID-19 between ancestral populations.

## Background

The coronavirus disease 2019 (COVID-19) pandemic has impacted the entire world, reshaping our day-to-day lives forever. To date, there have been a total of 774 million reported cases of COVID-19 with reported 7 million deaths from COVID-19 globally.^1^ In response to this pandemic, there have been attempts by the global scientific community to collect, categorize, and elucidate any information that could help with the prevention and treatment of this disease. Within these datasets, several risk factors have been identified, including, age, male sex, and past smoking status. However, one potential risk factor of COVID-19 infection that has not been as well characterized is ancestry.

Genetic ancestry is associated with different disease prevalence and COVID-19 is no exception. ^2–5^ Early on in the pandemic, it was noticed that people of different ancestries were impacted by COVID-19 differently.^6–8^ In 2020, Millet et al. investigated COVID-19 cases and death by across the United States of America, stratified by county. They found that COVID-19 disproportionately impacted African American communities after adjusting for age, poverty, and other comorbidities.^6^ Price-Haywood et al. investigated non-Hispanic African American patients and compared them to non-Hispanic European-ancestry patients in Louisiana. They found that even though the African American population accounted for 31% of the total cohort, this minority composed of 71% of the deaths. In response, there have been many studies that attempted to look at the underlying genetic differences to see if the difference in COVID-19 response was a genetic or socio-economic outcome.^9–15^ Shelton et al. found that ancestry was a hospitalization risk factor, however, they reported that the two genetic associations they found were responsible for this risk difference.^12^

One method to examine the biological response to COVID-19 infection is by investigating the circulating proteins levels of individuals who are acutely ill and comparing these levels to those in controls. This comparison can identify proteins, whose levels tend to be perturbed by COVID-19 infection. High-throughput oligonucleotide-aptamer protein measurement technology allows for the measurement of thousands of circulating proteins simultaneously from a single sample. In this study, samples were analyzed using the SomaScan assay (SomaLogic), an aptamer-based proteomics assay which utilizes chemically modified nucleotides, called SOMAmers (Slow Off-rate Modified Aptamers). The increased throughput of this technology allows for the measurement of many such proteins simultaneously. In spite of the benefits of proteomic analysis, there are only a few studies that looked at the proteomic differences between ancestries, and these ones focused primarily on the European ancestry.^14,16^ Previously, we found that an isoform of *OAS1* in people of European ancestry displayed a protective effect for COVID-19.^14^

Given the difference in rates of COVID-19 between different ancestries, it is important to assess whether these differences are biological in nature. However, there has been very little research as to how the proteome is impacted by COVID-19 stratified by ancestry. This study, using the Biobanque Québécoise de la COVID-19 (BQC19), assesses how each continental ancestry responds to COVID-19, as reflected by changes in circulating protein levels. The results of this study shed light on the host response to infection via changes to the proteome, and describes which responses differ by ancestry.

## Methods

### Overview of the study design

We used the SomaScan V4 assay to measure 4,979 circulating proteins in case and control samples from the BQC19, a biobank in Québec, Canada.^17^ We used linear mixed models (LMMs) to assess differences in protein levels between severe COVID-19 patients and controls who are hospitalized patients that either did not have COVID or presented with mild COVID symptoms, stratified by ancestry. The differences in the association between protein levels and COVID-19 severity was then compared between ancestries.

### Population

All samples were taken from the BQC19, this biobank recruited patients admitted to any of ten test sites in Quebec, Canada. We used genotype data provided by the BQC-19 to define the genetic ancestry of our populations and used phenotypic data provided by the BQC-19 to account for confounding factors, such as age, sex, and time since symptom onset.

### COVID-19 Case/Control Definitions

We used severe COVID-19 to define case status. Cases were defined as hospitalized individuals with COVID-19 as the primary reason for hospital admission, an RT-PCR confirmed COVID-19 status, and either died or required respiratory support beyond low-throughput nasal cannula. Controls were all individuals in the BQC-19 who did not meet this case definition. These controls were also admitted to the same hospitals within the participating hospital network in Quebec, Canada.^17^ This classification strategy has been previously used in the literature by the COVID-19 Host Genetics Initiative.^9,10,14,15^

### Ancestry Determination

Genetic ancestry was determined by a UMAP (v 0.2.8.0) projection of the BQC19 onto a reference set (Supp. Fig 1). This reference ancestral set was constructed by unrelated samples from the 1000 Genomes Project and the Human Genome Diversity Project. This reference set contained 117,221 SNPs that were available in the BQC19, had a minor allele frequency of >0.1%, and were LD-pruned (r^2^ < 0.8). The BQC-19 population was then projected on this reference and ancestry was assigned by a UMAP cluster assigned by hdbscan (v 0.8.27). This method was previously used by the COVID-19 HGI.^10,18^

### Protein measurements

We used the SomaScan (v4) platform to measure 5,284 circulating proteins from each sample. Of these 5,284 proteins, 305 non-human proteins were then removed from analysis using the same classification used previously in the literature.^19^ To adequately reflect the effects of COVID-19 infection, we limited our experiment to those biological samples procured within 30 days after symptom onset, with our linear mixed models accounting for the changes in protein levels over time. Blood plasma samples were collected in acid-citrate-dextrose tubes and frozen at −80 °C. Protein levels were measured using relative fluorescence units, and further normalized and calibrated by SomaLogic to remove any systematic bias. For the following statistical analysis, each protein sample across all patients was then normalized to have a mean of 0 and standard deviation of 1. This normalization was done prior to any ancestry stratification. The sample correction and proteomic normalization has been previously described in more detail by Butler-Laporte et al.^20^

### Statistical Analysis

To leverage the BQC19’s longitudinal samples, we used a linear mixed model (LMM) to group samples from the same patient using their BQC identifier. R packages, lme4 (1.1.35.1) and lmerTest (3.1.3) were used with R version 4.1.2 to generate these LMMs. This model further accounts for the patient’s age, sex and when the sample was taken relative to when the patient first had symptoms. These patients were further stratified by ancestry to examine how COVID-19 impacts protein levels by ancestry. The formula and sample code can be found in Supplementary file 1 and the GitHub (https://github.com/richardslab/BQC_Ancestry_Proteomics) respectively. A full table of all 4,979 proteins, their ancestry specific effect sizes, and their standard errors can be found in Additional File 1.

Each ancestry had all 4,979 proteins modelled in our linear mixed model. For each protein, severe COVID-19 illness had a reported effect size (beta) and p-value. This beta reports the difference between our case and control populations in each ancestry separately. These p-values were then corrected for multiple comparisons across all three models, resulting in a total of 14,937 tests with a false discovery rate of 5%.

To assess how COVID-19 impacts protein levels differently between ancestries we used both the beta and standard error from our LMM in each ancestry and derived a Z-score. A full formula can be found in Supplementary File 1.

These Z-scores were then converted into p-values based on a Chi-squared distribution. These p-values were then adjusted for multiple comparisons by applying a Benjamini-Hochberg correction, setting a false discovery rate of 5%. This test was chosen instead of using a heterogeneity test due to bias that can be introduced when using a small number of studies.^21^ We note that this threshold may be too liberal, given the relatively small sample sizes of non-European ancestry. Therefore, the results should be considered hypothesis generating.

## Results

### Sample Characteristics

Table 1 shows the total number of samples used in our study were separated by genetic ancestry. The samples were split into the six major ancestry groups as used by the HGI and the “other” category. These groups were: “European”, “African”, “East Asian”, “South Asian”, “Admixed American”, “Middle Eastern”, and “Other”. For the remainder of this study, we focused only on our three largest ancestral groups: European, African, and East Asian. For patient characteristics, see Supplementary Table 1.

**Table 1:** Sample characteristics in the BQC19.

|  | European | African | East Asian | Middle Eastern | Admixed American | South Asian | Admixed/ Other |
| --- | --- | --- | --- | --- | --- | --- | --- |
| Overall *n* (% of entire sample) | 797 (63%) | 196 (15%) | 79 (6%) | 76 (6%) | 51 (4%) | 28 (2%) | 45 (4%) |
| Median Age (IQR) | 70 (58 – 82) | 53 (43 – 65) | 56 (48 – 70) | 72 (54 – 80) | 54 (40 – 65) | 52 (38 – 62) | 52 (32 – 64) |
| # of Females (%) | 362 (45 %) | 98 (50 %) | 41 (52 %) | 28 (37%) | 22 (43%) | 10 (36%) | 22 (49%) |
| Severe COVID (%) | 223 (28%) | 72 (37%) | 27 (34%) | 31 (41%) | 19 (37%) | 7 (25%) | 5 (11%) |
| Median Days since Symptom Onset (IQR) | 9 (4 – 14) | 9 (5 – 15) | 9 (6 – 13) | 7 (2 – 13) | 12 (6 – 17) | 8 (4 – 13) | 8 (4 – 13) |

Numbers are presented as count (percentage) or (IQR). IQR is the interquartile range, which is the spread of the middle half of the distribution of the population.

### Different ancestries had different responses to COVID-19 infection

We first investigated how different ancestries protein levels responded to severe COVID-19 by stratifying the population using the following ancestries: European, East Asian, and African. In individuals of European ancestry 3,066/4,979 (62%) proteins had different (FDR < 0.05) proteins levels when comparing case and control populations (Figure 1A). Amongst individuals of African ancestry, 2,252/4,979 (45%) (Figure 1B) of the tested proteins had a difference in proteins levels between cases and control while 1,098/4,979 (22%) of the tested proteins in Asian ancestry individuals were classified as significantly different between our case and control populations. (Figure 1C). We note that the larger samples (European ancestry participants) had more different proteins than the smaller samples (African and Asian ancestry participants), which in part, reflects statistical power to detect such differences.

**Figure 1:**
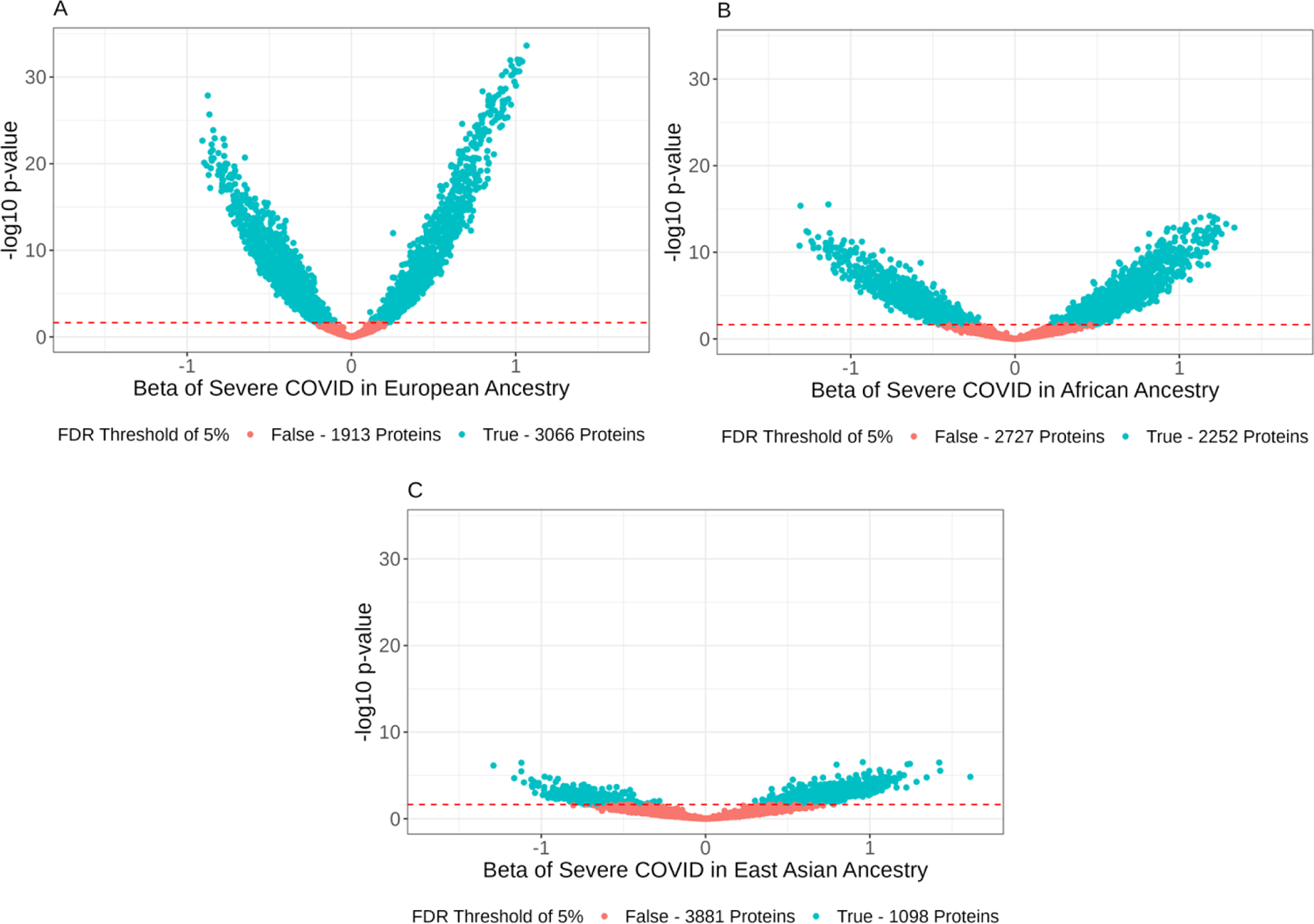
Volcano plots of the three tested ancestries of the BQC19: European – n = 797 (A), African – n = 196 (B), and East Asian – n = 79 (C).

The x-axis represents the effect size of the COVID variable in our linear mixed model. Our linear mixed model included age, sex, and days since symptom onset as co-variables, and used patient ID as a random factor to account for intra-patient correlation. All volcano plots are scaled to the same axis, each dot represents one tested protein. The colours representing True and False, indicate if the protein is significantly different between our case and control definition after multiple-comparison correction using the Benjamini-Hochberg method. The red dashed line represents our cutoff p = 0.021 to provide an FDR of 5%.

When comparing our European ancestries and our African ancestries, we saw that most proteins share the same trend upon severe COVID-19 infection. In Figure 2A, we compared all 4,979 of our tested protein’s European effect sizes to their African effect sizes. We found that they had a Pearson correlation of 0.868, 95% CI [0.861, 0.875]. We saw a similar, if weaker trend when comparing our European and East Asian ancestries with a respective Pearson correlation of 0.645, 95% CI [0.628, 0.661] (Figure 2B). Finally, we found that the trend continued between our East Asian and African ancestries in Figure 2C, with a Pearson correlation of 0.709, 95% CI [0.695, 0.722].

**Figure 2:**
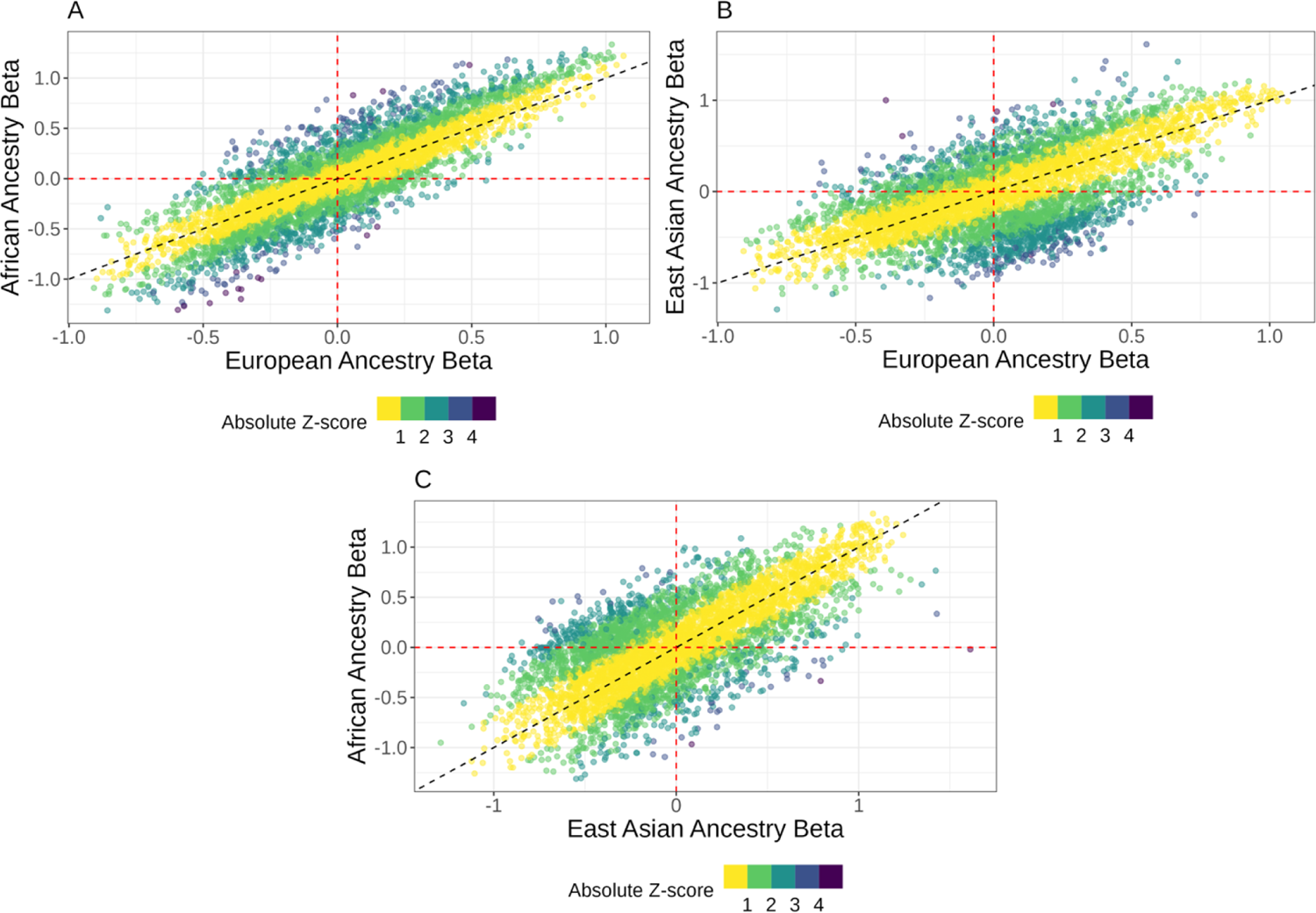
Pairwise Beta-Beta plots which displays all 4979 proteins and their associated COVID beta.

This effect size reflects the impact that COVID status has on the mean protein level in the tested ancestry. Each dot represents a single tested protein with its x-position and y-position representing the effect size per ancestry. In Figure 2A, the X position is the effect size associated with the European cohort and the Y position is the effect size in our African population. In Figure 2B, we are comparing the European and East Asian cohorts (x and y respectively), and in Figure 2C, we compare the East Asian (x-axis) and African effect sizes (y-axis). The red dashed lines indicate where an effect size of 0 would be and the black dashed line is the identity line, where a protein would have identical effect sizes in both ancestries.

These plots do not show any information on the standard error from these effect sizes. By colouring the plots based on their Z-score, we observed that the farther away from the dashed line – the identity line, the higher the Z-score. The proteins with Z-scores of 3.8 and higher are of interest for future investigation for precise targets. We noted that very few proteins show a marked deviation from the line of identity. Yellow is for proteins with the same response between ancestries while dark purple shows proteins that have significantly different responses to COVID between ancestries.

### Differences in how severe COVID-19 influences protein levels

We then calculated the ancestry-specific effect size of how COVID-19 status affected protein levels; we compared each ancestry in a pairwise fashion. Statistical significance of between-ancestry differences were determined using a Z-test (Methods). These p-values were then corrected for the 14,937 comparisons using an FDR – Benjamini-Hochberg – threshold of 5%. After applying this multiple testing correction, we found that 39 unique proteins had different betas across the three ancestries. 29 proteins had different COVID-19 responses between our European and African cohorts, 7 proteins had different COVID-19 responses between our European and East Asian cohorts, and 4 proteins had a different COVID-19 response between our East Asian and African cohorts (Supplemental Table 1).

As shown in Figure 3, many of the proteins discovered displayed effect sizes in the same direction but different magnitudes. Of note, in Figure 3A, 16 of the 29 proteins had effect sizes that were both negative and 6 proteins had effect sizes that were both positive – with the European counterpart consistently having the milder effect size, and the remaining seven proteins had effect sizes with opposing magnitudes. Of the seven proteins shown in Figure 3B, two proteins had COVID-19 responses in the same direction with different negative magnitudes, and the other five proteins had opposing effect sizes dependant on ancestry. Finally, in Figure 3C, all four proteins had opposing effect sizes between our East Asian and African populations.

**Figure 3:**
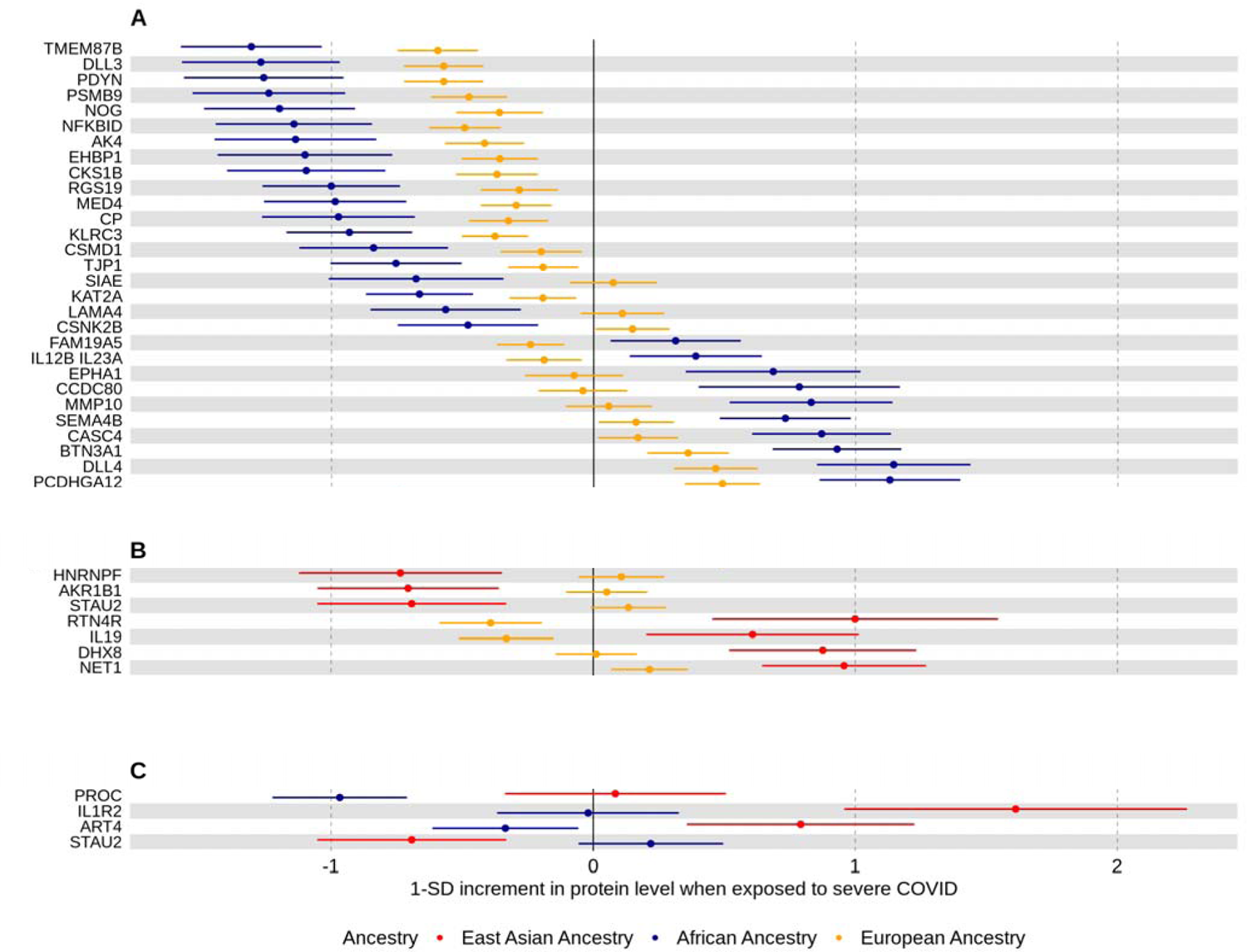
Forest plots of all proteins that were significantly different between two of the three ancestries tested.

Figure 3 shows Forest plots of the proteins that had a significant difference between ancestries (FDR < 0.05). Fig. 3A contains the 29 proteins that had significant differences in their effect sizes between our European and African ancestries and are displayed in order from lowest to highest African effect sizes. Of the 29 proteins, all were considered significantly different between case and control in the African ancestry while only 21 of them were considered to be significantly different from our control population in our European ancestry. Fig. 3B, contains the 7 proteins that passed our multiple correction threshold in our East Asian/European comparison is shown in order of their European effect sizes. Of the seven proteins, all seven had significant response to COVID-19 infection in our East Asian ancestry, while only three of the proteins showed a significant difference between our European case and control populations. Fig. 3C compares the East Asian and African effect sizes, in the order of lowest to highest African effect sizes. Two of the proteins were significantly different between our cases and controls in our African ancestry and three of our proteins had significant COVID-19 responses in our East Asian ancestry. In all plots the colour scheme is maintained: European (yellow), East Asian (Red), and African (blue). As in Figure 1, these changes are the effect sizes from the COVID variable of the linear mixed model. Error bars are the standard error of each effect size.

We noted that in all cases, when comparing differences in protein levels between two ancestries, the smaller of the two ancestries in sample size had the larger effect size. This may reflect the instability of estimating effect sizes in our smaller sample sizes, rather than true biological differences between the two groups.

### Validation of *OAS1* results

As previously mentioned, *OAS1* was identified by Zhou et al.^14^ to be a circulating protein that caused less severe COVID-19. Their conclusions hypothesized that COVID-19 infection would cause OAS1 levels to increase in European individuals. When we tested OAS1 protein levels, we found that they were increased in our severe COVID-19 patients in both European and African ancestries (unadjusted p-value of 1.0 × 10^-4^ and 2.7 × 10^-5^ respectively) (Figure 4).

**Figure 4:**
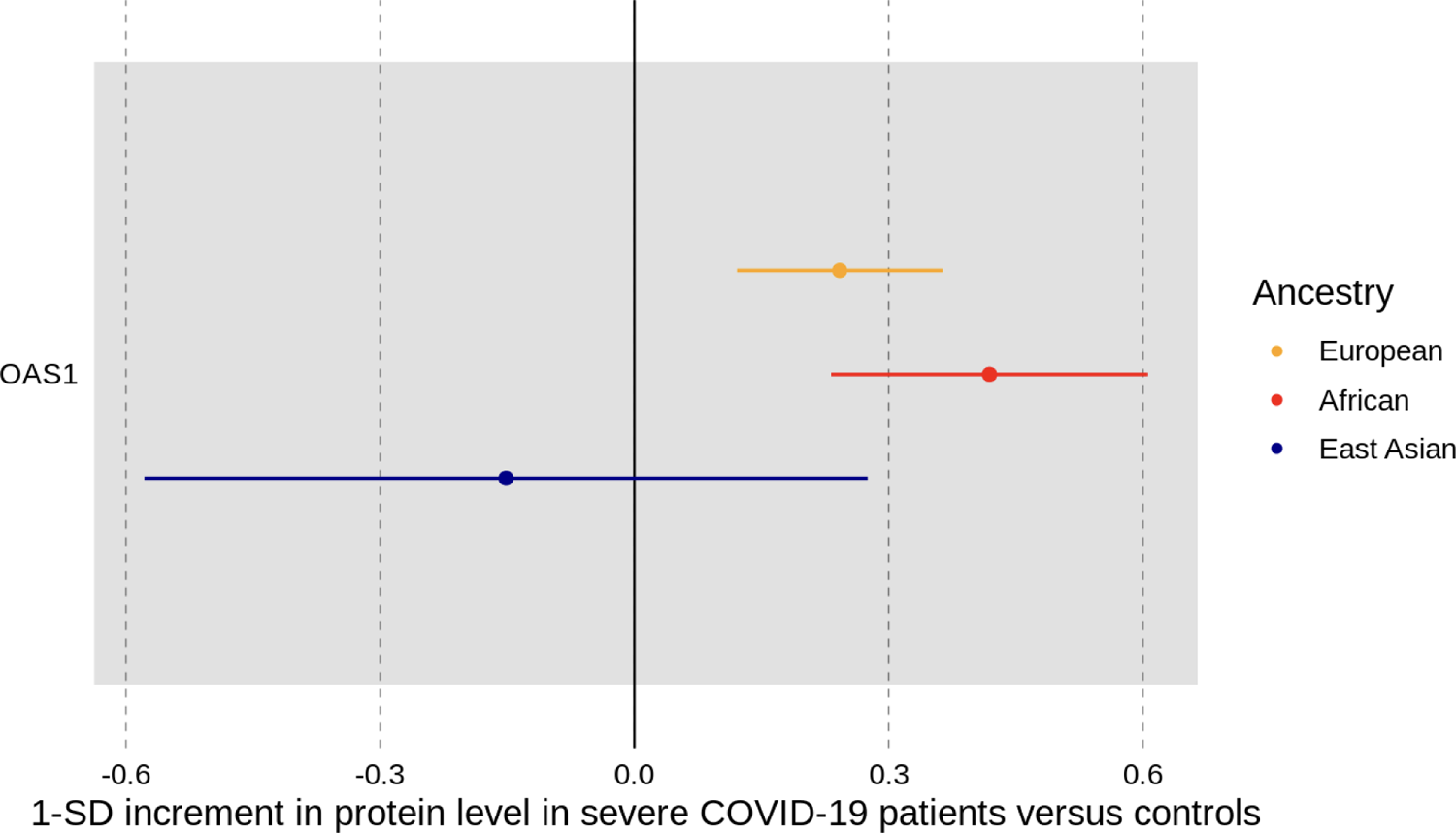
Forest plot of OAS1 when comparing case and control status across all three tested ancestries.

However, since the test in East Asian ancestry individuals was not well-powered due to the small sample size, there was no detectable difference in OAS1 levels between severe COVID-19 patients and controls (p-value of 0.49).

The effect size of each model is represented by the dot with the horizontal lines indicating the standard error associated with the effect size.

## Discussion

We leveraged the BQC19 to measure 4,979 circulating proteins across 1,272 samples from 909 patients. We then stratified these measurements by genetic ancestry and found 39 unique proteins had may have responded differently to severe COVID-19 infection. However, we note that these results should be viewed as hypothesis generating, and require replication, given the small sample sizes of non-European ancestries. Of these 39 proteins, we found that 29 showed a difference in their COVID-19 response between our European and African cohorts, 7 showed a difference between our European and East Asian cohorts, and 4 of had a difference between our East Asian and African cohorts – with one protein, STAU2 being found in two of our comparisons. Given the small sample sizes assessed in this study, and the large amount of data points being analyzed, replication of this work is essential to ensure the veracity of this data.

Our European – African ancestry comparison provided us with the most results most likely due to the stronger power of both models in comparison to our East Asian model. One of the proteins that displayed a significant difference between our European and African ancestries is FAM19A5. FAM19A5 is also known as TAFA5 and is in a family of homologous secreted proteins and acts as an adipokine.^24–26^ FAM19A5, unlike the rest of its family proteins, is expressed in both adipose tissue and the central nervous system which explains its presence as a circulatory protein. FAM19A5 has been shown to supress vascular smooth muscle cell proliferation and mice models with an overexpression of FAM19A5 have reduced scar tissue formation.^26^ Additionally, FAM19A5 has been shown to have an impact on hypothalamic inflammatory responses, with TNF-α administration increasing levels of hypothalamic FAM19A5 mRNA and when FAM19A5 itself was introduced to a mice model, the mice displayed a decreased food intake and increased body temperature.^27^ Considering that FAM19A5 was decreased in Europeans (effect size = −0.24, p = 2.6 × 10^-4^) and increased in our African population (effect size = 0.31, p = 0.015) this could be a factor in the severity of COVID-19 infection.

Of the four proteins that showed a difference between our East Asian and African ancestries, IL1R2 stood out as an interleukin receptor. IL1R2 is a decoy receptor which acts as a competitive binder of IL1B. The elevated levels of IL1R2 in our East Asian cohort upon COVID-19 infection should result in a lower impact of IL1B in the same cohort. Interestingly, IL1B was also measured directly by the SomaScan Assay, and we saw that it also had a differential response between East Asian and African populations (with elevated levels in our East Asian population in comparison to our African population), although this difference was not statistically significant after multiple testing correction (p-value=0.004). IL1B is a known marker of COVID-19 infection and is a part of the resultant cytokine storm.^28,29^ Furthermore, high levels of IL1B were shown to be associated with a reduced viral load of in the intestine of COVID-19 patients.^29^

There were seven proteins that showed a difference in their response to COVID-19 between our East Asian and European cohort. Of the seven proteins, STAU2 was the only protein found in both this comparison and the comparison between our East Asians and African cohorts. STAU2 is a double stranded RNA-binding protein that has been previously shown to interact with viral proteins such as NSP2 in COVID-19 and the Gag protein in HIV-1.^30,31^ Furthermore, STAU2 was shown to enhance the viral infectivity of HIV-1, with *STAU2* KO lines reducing the viral infectivity of HIV-1 by 55%.^30^ These results coupled with the results found here indicate that there might be a genetic difference in East Asians that should be further investigated to see if STAU2 also provides a similar protection to COVID-19 and to see if there is a similar interaction between HIV-1 infection and STAU2 protein levels in East Asian populations.

This study shows that while we should consider both the proteomic differences between different ancestries and their response to COVID-19, many of the circulating proteins respond in the same way regardless of ancestry. This study further shows that there are novel proteins that have not been explored due to the European ancestry-centric focus of many genomic, transcriptomic, and proteomic studies of COVID-19.

This study has important limitations. First, there was a relatively small number of individuals assessed in non-European ancestries. Since the European ancestry cohort has a much larger sample size, it is not surprising that the European ancestry individuals-based analyses yielded more findings due to the highest statistical power. More samples from our East Asian and African cohorts, as well as other ancestral groups that were omitted from this study due to their small sample sizes, would result in a stronger analysis. Potential confounders, such as smoking status and BMI, were not included in our analyses due to the large amount of missing data. It remains possible that the findings may not reflect direct causal effects of severe COVID-19 illness on circulating protein levels. Furthermore, we did not attempt to understand if any of the observed proteins had causal effects on COVID-19 outcomes. Another limitation of this study is that we only focused on how ancestries responded differently to a severe COVID-19 infection and did not investigate overall proteomic differences between ancestries or how these differences impact COVID-19 infection and a patient’s response of infection. Additionally, there likely exist proteomic differences unable to be captured by our measurement assay. Functional mutations may have affected the binding affinity of SOMAmers resulting in different levels of protein observed between ancestries if these protein-coding changes had different frequencies between the ancestries. Finally, we re-emphasize that replication of our results in diverse populations is necessary to better understanding trends linked with ancestry and proteomic responses.

## Conclusion

In summary, this work assessed how different genetic ancestries responded to severe COVID-19 infection by examining 4,979 circulating proteins. We found that while the majority of proteins responded in the same manner regardless of ancestry, there were 39 unique proteins that had suggestive differences in their response to COVID-19, which warrant functional follow-up studies.

## List of abbreviations

COVID-19: Coronavirus disease 19

BQC19: Biobanque Quebecois de la COVID-19

LMM: Linear mixed model

## Ethics Approval

Informed consent for inclusion in this study was obtained from every patient or their legal representatives. The Biobanque Québécoise de la COVID-19 (BQC19) received ethical approval from the IRB of the Jewish General Hospital and the Medical/Biomedical Committee of the Centre Hospitalier de l’Université de Montréal.

## Author Contributions

Conception and Design: TZ, YI, and JBR. Data Analyses: TZ. Data Acquisition: TN, GBL, DM, DA, LC. Data Verification: TZ, LC. Interpretation of the Data: TZ, YI, and JBR. Funding Acquisition: JBR. Methodology: TN, TL. Writing: TZ, YI, and JBR. Editing: TZ, TL, GBL, TN, and JBR. All authors gave final approval of the version to be published. The corresponding author attests that all listed authors meet authorship criteria and that no others meeting the criteria have been omitted. All authors read and approved the final manuscript.

## Data Availability

The BQC19 is an Open Science Biobank. Instructions on how to access data for individuals from the BQC19 is available at https://www.bqc19.ca/en/access-data-samples. To see the code used to analyze this data, it is available at https://github.com/richardslab/BQC_Ancestry_Proteomics.

## Acknowledgements

We would like to acknowledge the important contributions of research participants to help us better understand the causes and consequences of the COVID pandemic.

