## Supplemental Tables and Figures for "Differential circulating proteomic responses associated with ancestry during severe COVID-19 infection"

Supplementary Table 1: Unique patient characteristics in the BQC19, stratified by ancestry.

| | **European** | **African** | **East Asian** | **Middle Eastern** | **Admixed American** | **South Asian** | **Admix/ Other** |
|---|---|---|---|---|---|---|---|
| Overall *n* (%) | 584 (64%) | 147 (16%) | 53 (6%) | 46 (5%) | 33 (4%) | 19 (2%) | 27 (3%) |
| Median Age (IQR) | 73 (62 – 82) | 55 (43 – 66) | 58 (51 – 73) | 74 (54 – 82) | 56 (53 – 68) | 54 (38 – 66) | 54 (37 – 83) |
| # of Females (%) | 273 (47 %) | 75 (51 %) | 33 (61 %) | 22 (48%) | 13 (39%) | 7 (37%) | 15 (56%) |
| Severe COVID (%) | 175 (30%) | 61 (41%) | 19 (35%) | 17 (37%) | 13 (39%) | 5 (26%) | 1 (3.7%) |
| Median Days since Symptom Onset (IQR) | 10 (4 – 16) | 10 (7 – 16) | 9 (6 – 13) | 4 (1 – 14) | 16 (9 – 18) | 11 (5 – 17) | 9 (6 – 14) |

Numbers are presented as count (percentage) or (IQR). IQR is the interquartile range, which is the spread of the middle half of the distribution of the population.

### Supplemental Formula and Code

Formula

$$Z = \frac{\hat{b}_1 - \hat{b}_2}{\sqrt{SE_{\hat{b}_1}^2 + SE_{\hat{b}_2}^2}}$$

Z-test used to determine how different one ancestral model was from another. $\hat{b}_1$ and $\hat{b}_2$ are the estimated effect sizes of severe COVID-19 illness on protein level in models 1 and 2 respectively and $SE_{\hat{b}_1}$ and $SE_{\hat{b}_2}$ are for the standard errors of these respective estimates. $Z^2$ follows a chi-squared distribution with one degree of freedom.

LMM Formula:

Protein Level = $\beta_0$ + $\beta_1$ COVID + $\beta_2$ Age + $\beta_3$ Sex + $\beta_4$ DSO + (1 | BQCID)
DSO: Days since symptom onset
BQCID: Unique Patient ID which lets us track which samples come from each patient

Supplementary Figure 1: UMAP projection of the population of the BQC19

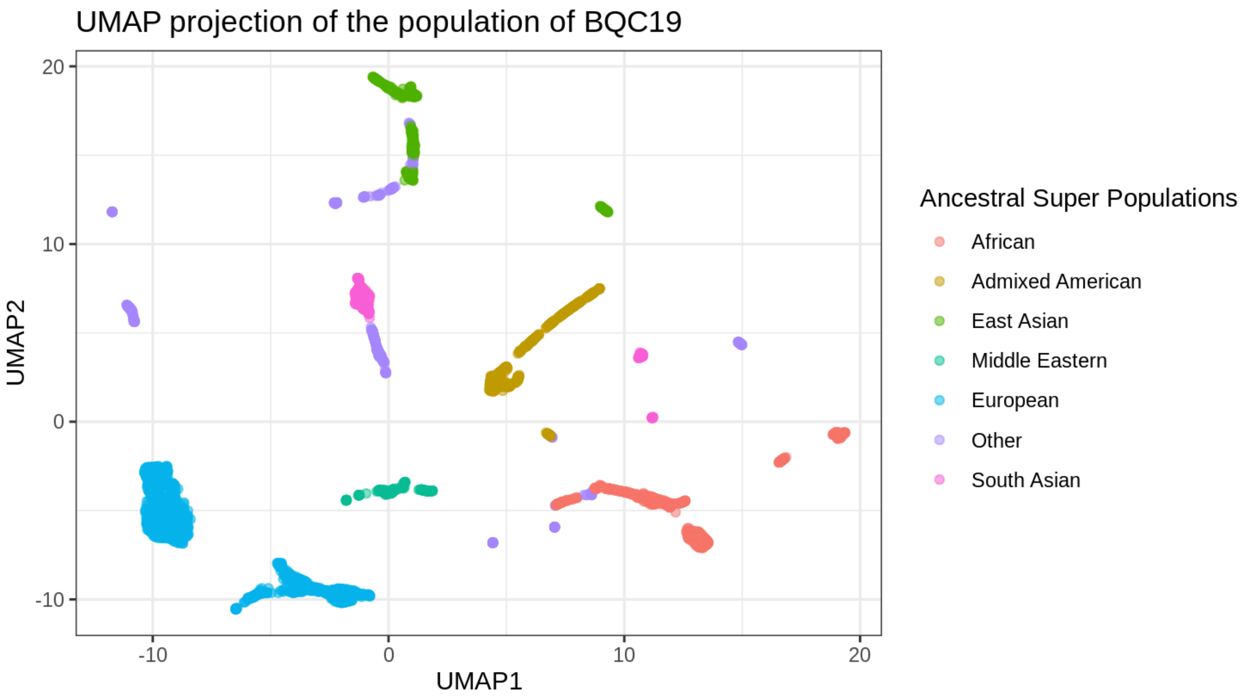


UMAP of all BQC19 tested patients sorted into the six super population as defined by HGI and an unclassified (“other”) population.
